# Immune response to SARS-CoV-2 variants of concern in vaccinated individuals

**DOI:** 10.1101/2021.03.08.21252958

**Authors:** Matthias Becker, Alex Dulovic, Daniel Junker, Natalia Ruetalo, Philipp D. Kaiser, Yudi T. Pinilla, Constanze Heinzel, Julia Haering, Bjoern Traenkle, Teresa R. Wagner, Mirjam Layer, Martin Mehrlaender, Valbona Mirakaj, Jana Held, Hannes Planatscher, Katja Schenke-Layland, Gérard Krause, Monika Strengert, Tamam Bakchoul, Karina Althaus, Rolf Fendel, Andrea Kreidenweiss, Michael Koeppen, Ulrich Rothbauer, Michael Schindler, Nicole Schneiderhan-Marra

**Author notes:** these authors contributed equally to this work (Matthias Becker, Alex Dulovic). Contact Information Nicole Schneiderhan-Marra –. Postal Address – Markwiesenstrasse 55, 72770 Reutlingen, Germany. Michael Schindler –. Email Address Postal Address – Elfriede-Aulhorn-Strasse 6, 72076 Tübingen, Germany. Ulrich Rothbauer –. Email Address Postal Address – Markwiesenstrasse 55, 72770 Reutlingen, Germany. **Competing Interests** T.R.W., P.K., N.S.M. and U.R. are named as inventors on a patent application (EP 20 197 031.6) claiming the use of the described Nanobodies used in the NeutrobodyPlex for diagnosis and therapeutics filed by the Natural and Medical Sciences Institute. The other authors declare no competing interest.

## Abstract

The SARS-CoV-2 pandemic virus is consistently evolving with mutations within the receptor binding domain (RBD)^1^ being of particular concern^2-4^. To date, there is little research into protection offered following vaccination or infection against RBD mutants in emerging variants of concern (UK^3^, South African^5^, Mink^6^ and Southern California^7^). To investigate this, serum and saliva samples were obtained from groups of vaccinated (Pfizer BNT-162b2^8^), infected and uninfected individuals. Antibody responses among groups, including salivary antibody response and antibody binding to RBD mutant strains were examined. The neutralization capacity of the antibody response against a patient-isolated South African variant was tested by viral neutralization tests and further verified by an ACE2 competition assay. We found that humoral responses in vaccinated individuals showed a robust response after the second dose. Interestingly, IgG antibodies were detected in large titers in the saliva of vaccinated subjects. Antibody responses showed considerable differences in binding to RBD mutants in emerging variants of concern. A substantial reduction in RBD binding and neutralization was detected for the South African variant. Taken together our data reinforces the importance of administering the second dose of Pfizer BNT-162b2 to acquire high levels of neutralizing antibodies. High antibody titers in saliva suggest that vaccinated individuals may have reduced transmission potential. Substantially reduced neutralization for the South African variant highlights importance of surveillance strategies to detect new variants and targeting these in future vaccines.

## Introduction

Since the initial outbreak in Wuhan, China in late 2019^9,10^, SARS-CoV-2 has evolved into a global pandemic, with more than 112 million infections and 2.5 million deaths (as of February 26^th^, 2021)^11^, impacting severely on mental health^12,13^ and global economics^14^. In response, the scientific community has made unprecedented progress, resulting in the generation of multiple vaccines, using a variety of different approaches^8,15,16^, such as the Pfizer BNT-162b2 vaccine, which encodes a full-length trimerized spike protein^17^. In parallel, SARS-CoV-2 is continually evolving which might impact its infectivity^2^, transmission^3,18,19^ and viral immune evasion^20,21^. To date, advanced genomic approaches have identified thousands of variants of SARS-CoV-2 with multiple RBD mutations circulating due to natural selection^4,22^. The variability of RBD epitopes is of specific concern as such mutations might reduce vaccine efficacy, increase viral transmission or impair acquired immunity by neutralizing antibodies^2,23,24^. For the pandemic to be brought under control, herd immunity must be achieved through vaccination. However, there is discourse about how long antibodies generated during the first wave persist, with some studies suggesting seroreversion between two to three months^25^, while others find antibodies present for up to seven or eight months post-infection^26-28^. Alarmingly, antibodies generated during the first wave also appear to have reduced immunoreactivity and neutralization potency towards emerging variants^27^.

As the virus is known to continually mutate, particularly the emerging UK (B.1.1.7)^3^, South African (B.1.351)^5^, Brazil (P1)^29^, Mink (Cluster 5)^6^ and Southern California (hereon referred to as “LA” (B1.429)^7^ variants are of concern. The UK variant may have an increased risk of transmission^30^ and potentially increased mortality^18,30^. It further exhibits reduced neutralization susceptibility^21,31^, which is most substantially related to a subset of RBD-specific monoclonal antibodies^31^. The N501Y mutation appears to mediate increased ACE2-RBD interaction^1^ and is known to be critical for SARS-CoV-2 infection *in vivo* in mice^32^. Similarly, the South African variant which is now spreading globally, has two escape mutations within the RBD (K417N and E484K)^5^ in addition to the N501Y mutation. The combination of these three point mutations might result in both a higher infection rate and reduced capacity of neutralizing antibodies produced against variants without RBD-mutations of concern (hereon referred to as “wild-type”). In light of these developments, and in spite of increasing data provided by vaccine companies, it remains unclear whether vaccines formulated against the original Wuhan strain of the virus will remain effective against new and emerging variants such as UK or South Africa. To understand this, we characterized the antibody response post vaccination with the Pfizer BNT-162b2 vaccine in both serum and saliva and then investigated the presence and efficacy of neutralizing antibodies against emerging variants of concern (UK, South Africa, Mink and LA).

## Results

To analyze the humoral response generated by vaccination, SARS-CoV-2 reactive antibody titers in serum samples from vaccinated, convalescent (hereon referred to as “infected”) and un-infected (hereon referred to as “negative”) individuals were measured using MULTICOV-AB^33^ (**Fig. 1**). Descriptions of all groups of donors can be found in **Extended Data Table 1**. Vaccinated individuals had not been previously infected with SARS-CoV-2 as demonstrated by the absence of anti-Nucleocapsid IgG and IgA (**Fig. 1a**). As expected, there was typical variation in antibody titers reflecting individual immune responses (**Fig. 1a and b**). When comparing between vaccine doses (**Fig. 1c and d**), all vaccinated subjects showed an enhanced antibody response with increasing time after the first dose and a further significant boost after the second dose. This boosting effect was so pronounced that it reached the upper limit of detection for MULTICOV-AB, as confirmed by a dilution series (**Extended Data Fig. 1**).

**Figure 1.**
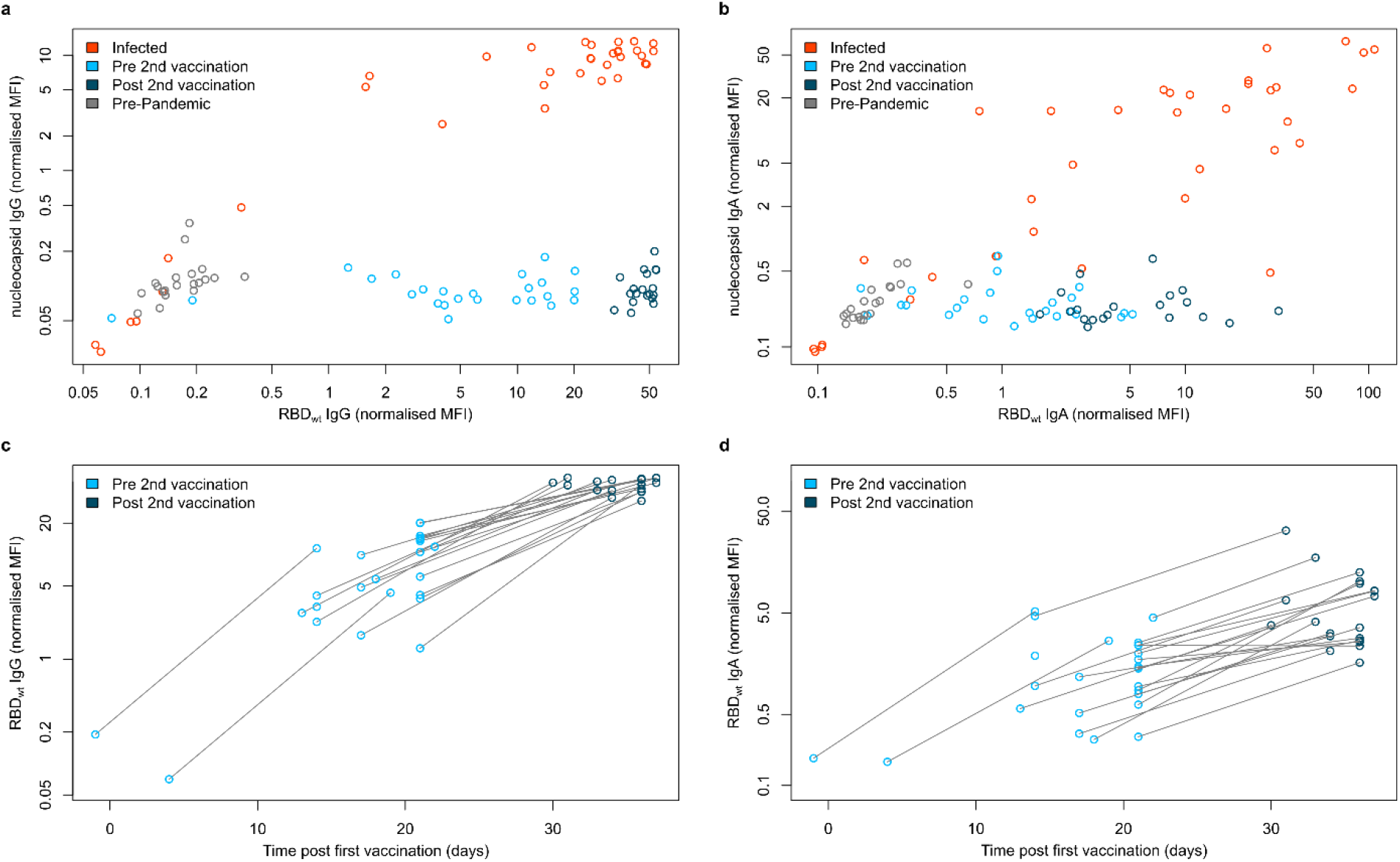
IgG and IgA response in serum samples of vaccinated, infected and negative individuals. IgG (**a** and **c**) and IgA (**b** and **d**) response in sera from vaccinated (pre second vaccination (light blue, N=25), post second vaccination (dark blue, N=20)), infected (red) (n=35) and negative (gray) (n=20) individuals was measured with MULTICOV-AB. IgG (**a**) and IgA (**b**) response is shown as normalized RBD versus normalized Nucleocapsid MFI values allowing for visualization of separation between the different groups. Increasing antibody titers in vaccinated individuals for IgG (**c**) and IgA (**d**) is shown with increasing days post vaccination, with samples colored based on whether they are before (light blue) or after (dark blue) the second dose. Lines indicate paired samples from the same donor.

To expand our understanding of the immune response of vaccinated individuals, we analyzed their saliva for IgA and IgG antibodies. The saliva of infected and negative individuals served as controls. Infected individuals had significantly higher levels of IgA than negative (p-value 0.0008) or vaccinated individuals (p-value 0.03), with no significant difference seen between vaccinated and negative individuals (p-value 0.23) (**Fig. 2a**). Conversely, the IgG response in saliva of vaccinated individuals was significantly higher than either infected (p-value <0.0001) or negative individuals (p-value <0.0001) (**Fig. 2b**). These results were verified using a second antibody test measuring IgG in saliva (**Extended Data Fig. 2**). We also identified that vaccination with Pfizer BNT-162b2 does not appear to offer any cross-protection against other endemic coronaviruses (**Extended Data Fig. 3**), as seen by absence of change in antibody titers following vaccination.

**Figure 2.**
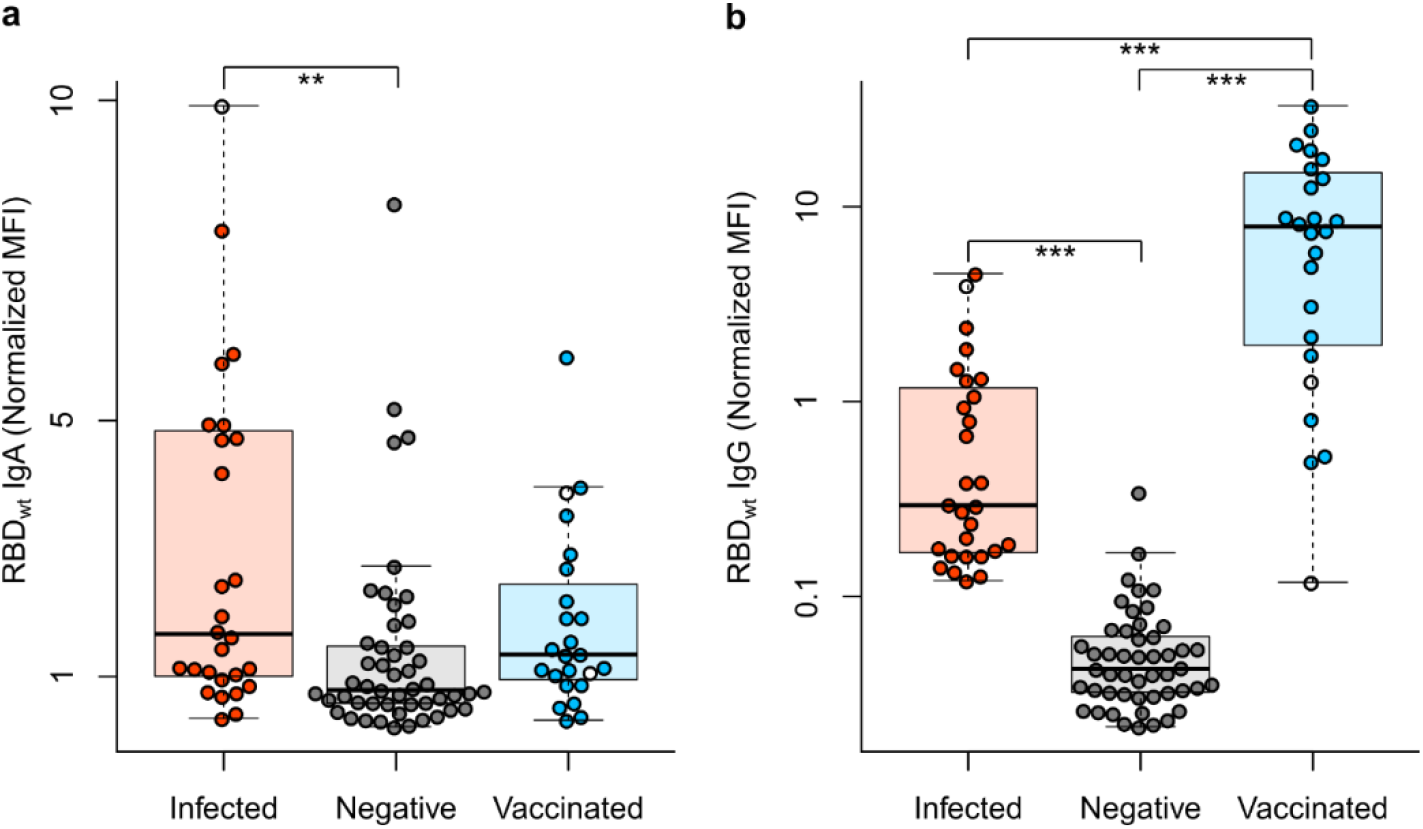
IgA and IgG response in saliva samples of vaccinated, infected and negative individuals. Box and whisker plots (1.5 IQR) for the IgA (**a**) and IgG (**b**) response in saliva of vaccinated (n=22), infected (n=26) and negative (n=45) individuals. All samples were measured three times using MULTICOV-AB, normalized against QC values to remove confounding effects, and the mean calculated and displayed. Panel **b** is presented using a logarithmic scale for clarity. As additional controls, one infected and then vaccinated sample and two vaccinated samples from individuals not in contact with active COVID-19 infections are displayed as clear circles. Statistical significance was calculated by Mann-Whitney-U with significance determined as being <0.01. **<0.001 ***<0.0001

Having determined the humoral response induced by Pfizer BNT-162b2, we then examined how emerging RBD mutations present in different variants of concern impact antibody binding. For this, we included RBD mutants for the UK (501Y), South African (417N, 484K and 510Y), Mink (453F) and LA (452R) variants in MULTICOV-AB. For the UK variant, a nearly identical antibody response was observed for vaccinated and infected individuals compared with wild-type variant (Kendall’s tau 0.965) (**Fig. 3a**). In contrast, a varied and reduced immune response was visible for the South African variant in both groups (Kendall’s tau 0.844) (**Fig. 3b**). Both the Mink and LA variants had a similar response as the wild-type variant (**Extended Data Fig. 4**). Having seen that antibody binding responses were reduced in the context of RBD mutants in the South African variant, we examined its neutralizing potential on samples from vaccinated individuals using a virus neutralization test (VNT)^34^, employing a patient-derived South African variant of the SARS-CoV-2 virus. Despite detectable variation, the VNT revealed substantially reduced neutralization for the South African variant for sera obtained from vaccinated and infected individuals (**Fig 4a**). We further confirmed these findings using an ACE2 inhibition assay (**Fig. 4b**). Here, we additionally observed increased neutralization capacities for both wild-type (**Fig. 4c**) and the South African variant (**Fig. 4d**), in all samples derived from vaccinated individuals following the second dose, which was further confirmed by the NeutrobodyPlex ^35^ (**Extended Data Fig. 5**). Overall, our results showed individual differences in neutralization capacity for RBD mutations found in variants (**Extended Data Fig 6**), with minimal to no change in neutralization for the UK, Mink or LA variants. Conversely, neutralization capacity versus the South African variant was severely compromised.

**Figure 3.**
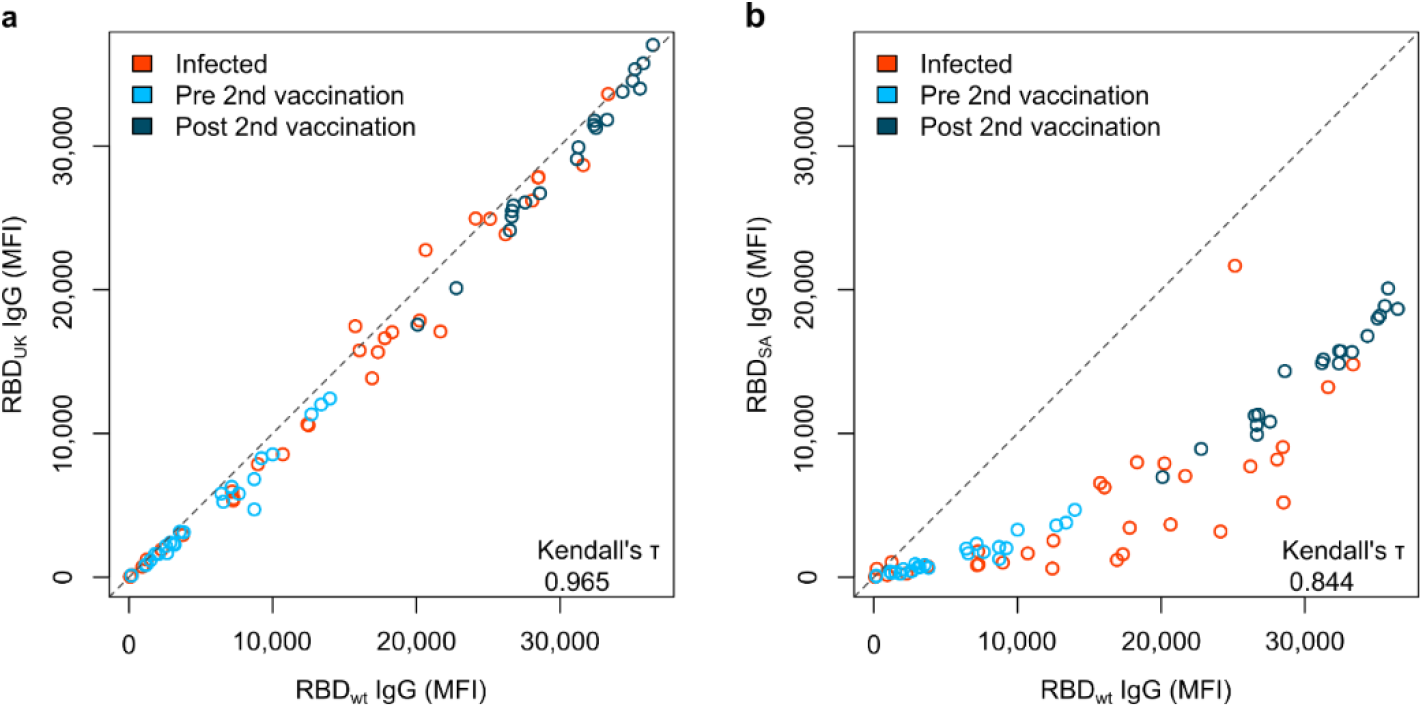
South African RBD mutant has a reduced response compared to UK RBD mutant. UK (**a**) and South African (SA) RBD mutants (**b**) have differing effects upon antibody binding. RBD mutant antigens were generated and added to MULTICOV-AB to measure the immune response towards them in sera from vaccinated pre-second dose (light blue) (n=25), post-second dose (dark blue) (n=20) and infected (red) (n=35) individuals, compared to the wild-type (wt) RBD. A linear curve (y=x) is shown as a dashed grey-line to indicate identical response between wild-type and mutant. Kendall’s tau was calculated to measure ordinal association between the mutant and wild-type.

**Figure 4.**
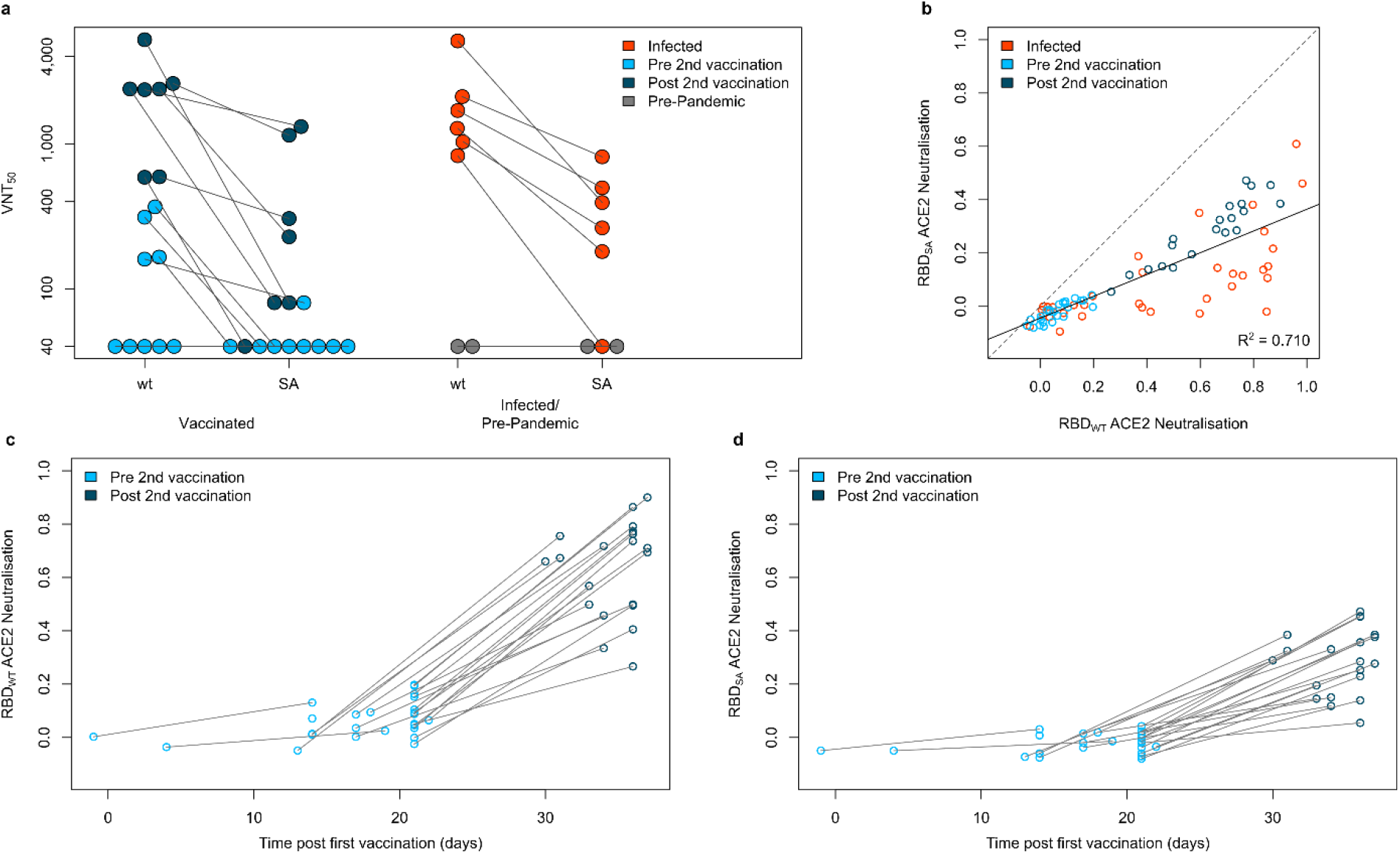
South African RBD variant has decreased neutralization compared to wild-type in vaccinated and infected samples. Neutralization for the South African variant (SA) displayed as virus neutralizing titers (VNT_50_) was measured in a virus neutralization assay compared to a wild-type variant (wt) (**a**) with sera from vaccinated (pre second vaccination (light blue, N=9), post second vaccination (dark blue, N=7)), infected (red, N=6) and negative (pre-pandemic) (grey, N=2) individuals. To confirm the reduction in neutralization seen, an ACE2 competition assay was developed and used to measure neutralization capacity for wild-type RBD (wt) and the South African RBD mutant (SA) (**b**) on sera from vaccinated (pre second vaccination (light blue, N=25), post second vaccination (dark blue, N=20)), infected (red, N=35) and negative (pre-pandemic, grey, N=20) individuals. 0 indicates that no neutralization is present while 1 indicates maximum neutralization. A linear regression (y = −0.044 + 0.408x) for all samples is shown in gray with the R^2^ included. When examining vaccinated samples only, wild-type neutralization (**c**) is significantly increased following the second vaccine dose. For South African neutralization (**d**), while it is increased following the second dose, there is significant reduction when compared to wild-type. Lines in (**a**), (**c**) and (**d**) indicate paired samples from the same donor.

## Discussion

RBD mutants are particularly important to track and study due to the role of the RBD:ACE2 interaction site in virus transmission and neutralization^1,18^ and their potentially increased infectivity^3^ or lethality^30,36^. This tracking has led to the identification of several variants of concern, notably the UK^3^, South African^5^, Mink^6^ and LA^7^ variants. We initiated this study to reveal the vaccine-induced immune response and most importantly to shed light on the still controversial question of how efficiently antibodies can bind and neutralize SARS-CoV-2 variants of concern. The presence of large titers of IgG antibodies within the saliva of vaccinated individuals far exceeded those seen in convalescent individuals. This was both surprising and welcome as it could indicate that vaccination might confer a sterilizing immune response in the oral cavity and thereby lower virus transmission. Focusing on antibody response, we examined in detail the effects of RBD mutations observed in emerging variants of concern. While only minor differences were detectable for the UK, Mink and LA variants, a substantial reduction in RBD binding antibodies was observed for the South African variant. These findings were confirmed at a functional level by a VNT using patient-derived viral isolates, which showed a significant decrease in the neutralizing capacity of sera from vaccinated or infected individuals for the South African variant. This could provide a reasonable explanation for infection in vaccinated or convalescent individuals with the South African variant and suggests a potential reduction in efficacy of the Pfizer BNT-162b2 vaccine from a B cell perspective.

This study is limited by the sample size, the restriction to only two time points after vaccination for data analysis, and the lack of paired saliva and serum samples for our infected and negative groups. However, we want to note that our sample size examined is similar to or larger than most other sample sets used to examine immune response to mutants in detail ^37^, and that we examined paired serum and saliva samples for all of our vaccinated subjects. Furthermore, we performed VNT assays comparing wild-type to the South African variant with authentic virus isolates on human cells, in contrast to utilizing a pseudotype neutralization assay^37^ or genetically engineered wild-type variant^38^. Notably, we focused our study on a detailed characterization of the humoral immune response, as a proxy of an individual’s immune response, as T-cell immunity has already been extensively studied^26,39^. Future work should investigate the antibody response and their persistence from different vaccines (i.e. AstraZeneca-Oxford, Moderna) against similar or newly emerging RBD mutants over a longer timeframe with increased sample size.

Viewed in a larger context, the impaired RBD-binding capacity to mutations in emerging variants of concern highlights the importance of updating current vaccines accordingly. Particular attention should be paid to the South African variant, because of the reduced neutralizing potency identified, while spontaneous independent enrichments of the E484K mutation have been observed in several countries^40^.

## Methods

### Data Reporting

No statistical analysis was used to determine sample size. The experiments were not randomized and the investigators were not blinded during either experimentation or data analysis

### Study recruitment, sample collection and ethics statement

Serum and saliva samples were collected from healthcare workers, vaccinated with the Pfizer BNT-162b2 vaccine. Two serum samples were collected from each individual. The first collection took place on average 17 days following administration of the first dose, while the second collection took place on average 12 days following administration of the second dose. Saliva samples were collected 7-10 days following administration of the second dose. Infected serum and saliva samples were collected from different groups of individuals. Serum samples were collected from individuals hospitalized at Universität Klinikum Tübingen between 25.03.2020 and 22.01.2021. All individuals tested positive for SARS-CoV-2 by PCR. Saliva samples were collected from individuals who had previously been infected with SARS-CoV-2 between 07.03.2020 and 19.06.2020. All individuals had previously tested positive by PCR, or were confirmed as being previously infected by ELISA measurements plus the presence of at least one key symptom (i.e. Coughing, Fever). Negative serum and saliva samples were also from different groups of individuals. Negative serum samples were purchased from Central Biohub. All of these samples were collected pre-pandemic. Negative saliva samples were collected at the Institute of Tropical Medicine, Universität Tübingen from negative individuals. As additional controls, serum and saliva samples were collected from two individuals vaccinated with Pfizer BNT-162b2, who do not have contact with active SARS-CoV-2 infected patients, and one individual who had been previously infected with SARS-CoV-2 and was later vaccinated.

For serum collection, blood was taken by venipuncture, the serum extracted and then frozen at −80°C until use. For saliva collection, all individuals spat directly into a collecting tube. To inactive the samples, TnBP and Triton X-100 were added to final concentrations of 0.3% and 1X respectively. Saliva samples were then frozen at −80°C until further use.

This study was approved the Ethics Committee of Eberhard Karls University and the University Hospital Tübingen under the approval number 312/2020BO1 (Coro-Buddy) to Dr. Andrea Kreidenweiss, Institute for Tropical Medicine, University Hospital Tübingen and Eberhard Karls University Tübingen, and 312/2020BO2 to Dr. Karina Althaus, Institute for Clinical and Experimental Transfusion Medicine, University Hospital Tübingen. All participants gave written informed consent. Characteristics of vaccinated and infected serum donors can be found in **Extended Data Table 1**. Characteristics of vaccinated, infected and negative saliva donors can be found in **Extended Data Table 2**.

### Expression of RBD Mutants

The pCAGGS plasmid encoding the receptor binding domain (RBD) of SARS-CoV-2 was kindly provided by F. Krammer ^41^. RBDs of SARS-CoV-2 variants of concern were generated by PCR amplification of fragments from wildtype DNA template followed by fusion PCRs to introduce described mutation N501Y for the UK variant and additional mutations K417N and E484K for the South African variant ^36,42,43^. Forward primer RBDfor 5′- ATA TCT AGA GCC ACC ATG TTC GTG TTT CTG G - 3′ and reverse primer N501Yrev 5′- CCA CGC CAT ATG TGG GCT GAA AGC CGT AG - 3′ were used for amplification of fragment 1, forward primer N501Yfor 5′- GGC TTT CAG CCC ACA TAT GGC GTG GGC TAT CAG C - 3′ and reverse primer RBDrev 5′- AAG ATC TGC TAG CTC GAG TCG C - 3′ were used for amplification of fragment 2. Both fragments containing an overlap sequence at the 3’ and 5’ end were fused by an additional PCR using forward primer RBDfor and RBDrev. Based on cDNA for the UK variant, additional mutations of the South African Variant were introduced by PCR amplification of 3 fragments using forward primer RBD-for and reverse primer K417Nrev 5′- GTT GTA GTC

GGC GAT GTT GCC TGT CTG TCC AGG G - 3′, forward primer K417Nfor 5′- GAC AGA CAG GCA ACA TCG CCG ACT ACA ACT ACA AGC - 3′ and reverse primer E484Krev 5′- GCA GTT GAA GCC TTT CAC GCC GTT ACA AGG GGT - 3′, forward primer E484Kfor 5′- GTA ACG GCG TGA AAG GCT TCA ACT GCT ACT TCC C - 3′ and reverse primer RBDrev. Amplified fragments were assembled by subsequent fusion PCR using forward primer RBDfor and RBDrev. RBD mutation L452R as recently reported for the SARS-CoV-2 variant of concern identified in Southern California (referred to in this manuscript as “LA”), was introduced using primer RBD-for and reverse primer L452Rrev 5′- CGG TAC CGG TAA TTG TAG TTG CCG CCG - 3′ for amplification of fragment 1 and forward primer L452Rfor 5′- GGC AAC TAC AAT TAC CGG TAC CGG CTG TTC CGG AAG - 3′ for fragment 2. Both fragments were subsequently fused using primers RBDfor and RBDrev. DNA coding for mutant RBDs (amino acids 319-541 of respective spike proteins) were cloned into Esp3I and EcoRI site of pCDNA3.4 expression vector with N-terminal signal peptide (MGWTLVFLFLLSVTAGVHS) for secretory pathway that comprises Esp3I site. All expression constructs were verified by sequence analysis.

### Bead coupling

Coupling of RBD mutant antigens was done by Anteo coupling (#A-LMPAKMM-10, Anteo Tech Reagents) following the manufacturer’s instructions. Briefly, 100 μL of spectrally distinct populations of MagPlex beads (1.25×10^6^) (Luminex) were activated in 100 μL of AnteoBind Activation Reagent for 1 hour at room temperature. The activated beads were washed twice with 100 μL of Coupling Buffer using a magnetic separator. Following this, 50 μg/mL of antigen (diluted in Coupling buffer) was added to the beads and incubated for 1 hour at room temperature. The beads were then washed twice with 100 μL Coupling buffer and blocked for 1 hour at room temperature in 0.1% BSA in Coupling Buffer. After washing twice in storage buffer, the beads were stored at 4°C until further use.

### MULTICOV-AB

MULTICOV-AB, a multiplex immunoassay which simultaneously analyses 20 antigens was performed as previously described ^33^ on all samples. In addition to the antigens presently included in MULTICOV-AB (**Extended Data Table 3**), RBD mutants from variants of concern were also included for measurements of all serum samples. All RBD mutants except the Mink variant (#40592-V08H80, Sino Biological) were produced in-house. Both IgG and IgA were measured for all serum samples. To adapt MULTICOV-AB to analyze antibodies in saliva, the dilution factor was changed from 1:400 for serum to 1:6 for saliva. Saliva samples were diluted into assay buffer inside a sterile workbench. 25 μL of diluted sample was then added to 25 μL of 1X Bead Mix ^33^ using a 96-well plate (#3600, Corning). Samples were incubated for 2 hours at 20°C, 750 rpm on a Thermomixer (Eppendorf), after which the unbound antibodies were removed by washing three times with Wash Buffer (1x PBS, 0.05% Tween-20) using a microplate washer (Biotek 405TS, Biotek Instruments GmBH). Bound antibodies were detected using either 3 μg/mL RPE-huIgG (#109-116-098, BIOZOL) or 5 μg/mL RPE-huIgA (#109-115-011, BIOZOL) by incubation for 45 mins at 20°C, 750 rpm on a Thermomixer. Following another washing step, beads were re-suspended in 100 μL of Wash Buffer and re-shaken for 3 mins at 20°C, 1000 rpm. Plates were then measured using a FLEXMAP3D instrument (Luminex) using the same settings as ^33^. Each sample was measured in three independent experiments. Raw median fluorescence intensity (MFI) values were divided by the MFI of QC samples included on each plate to produce a normalization value.

### Saliva IgG ELISA

To validate saliva measurements by MULTICOV-AB, samples were re-measured using an in-house ELISA established by the Institute of Tropical Medicine, Universität Tübingen. Saliva samples were analyzed for SARS-CoV-2 wild-type RBD reactive IgG antibodies by an in-house ELISA developed at the Institute of Tropical Medicine, University of Tübingen. SARS-CoV-2 RBD recombinant protein was dissolved in PBS to a final concentration of 2 µg/mL. 50 µL was then coated into 96well Costar microtiter high binding plates (#3590, Corning) and blocked at 4°C overnight with The Blocking Solution (Candor Bioscience GmBH), at room temperature on a microplate shaker set to 700 rpm. Before each of the following steps, wells were washed with PBS/ 0.1% Tween20. Saliva samples were diluted using The Blocking Solution (1:3 – 1:729) and 100 μL added to each well. Plates were then incubated at room temperature for 1 hour. For detection, 1:20,000 biotinylated anti-human IgG (#109-065-008, Jackson Immuno Research Laboratories) and 1:20,000 Streptavidin-HRP (#109-035-098) were added and incubated for 1 hour and 30 mins respectively. For visualization, TMB was added and the reaction was stopped using 1M HCl. The plate was read at 450 nm and 620 nm using a microplate reader (CLARIOstar, BMG LABTECH). Data is presented as concentration in ng/mL as estimated by a respective dilution series of highly pure human IgG (#31154, ThermoFisher).

### Neutralization Assays

Viral neutralization assays for the wild-type (Tü1) variant and the South African variant were performed as previously described ^34^ on 16 vaccinated, 6 infected and 2 negative serum samples. Briefly, Caco-2 cells were cultured at 37°C with 5% CO_2_ in Dulbecco’s modified Eagle medium (DMEM), supplemented with 10% fetal calf serum (FCS), 2 mM L-glutamine, 100 mg/mL penicillin-streptomycin and 1% non-essential amino acids (NEAA). The clinical isolate (200325_Tü1) ^34^ which belongs to the lineage B.1.126 is referred to as “wild-type” in this manuscript. The South African variant (210211_SaV) was isolated from a throat swab collected in January 2021 at the Institute for Medical Virology and Epidemiology of Viral Diseases, University Hospital Tübingen, from a PCR-positive patient. 100 µL of patient material was diluted in medium and used to directly inoculate 150,000 Caco-2 cells in a 6-well plate. At 48 hours post-infection, the supernatant was collected, centrifuged and stored at −80°C. After two consecutive passages, the supernatant was tested by qRT-PCR confirming the presence of three point mutations (N501Y, K417N and E484K). NGS confirmed that the clinical isolate belongs to the lineage B.1.351.Caco-2 cell infection with 210211_SAv was detected by Western blotting, using sera from a convalescent patient. Multiplicity of infection determination (MOI) was conducted by titration using serial dilutions of both virus stocks. The number of infectious virus particles per millimeter was calculated as (MOI x cell number)/(infection volume), where MOI = -ln (1 – infection rate). For neutralization experiments, 1×10^4^ Caco-2 cells/well were seeded in 96-well plates the day before infection in medium containing 5% FCS. Cells were co-incubated with SARS-CoV-2 clinical isolate 200325_Tü1 or SARS-CoV-2 clinical isolate 210211_SAv at an MOI of 0.7. Patient sera were added in serial 2-fold dilutions from 1:40 – 1:5,120. At 48 hours post infection, cells were fixed with 80% acetone for 5 mins, washed with PBS, and blocked for 1 hour at room temperature (RT) with 10% normal goat serum (NGS). Cells were incubated for 1 hour at RT with 100 µL of serum from a hospitalized convalescent donor in a 1:10,000 dilution and washed 3 times with PBS. 100 µL of goat anti-human Alexa594 (1:2,000) in PBS was used as a secondary antibody for 1 hour at RT. Cells were then washed 3 times with PBS and counterstained with 1:20,000 DAPI solution (2 mg/mL) for 10 mins at RT. For quantification of infection rates, images were taken with the Cytation3 (BioTek) and DAPI-positive and Alexa 594-positive cells were automatically counted by the Gen5 software (BioTek). Virus neutralizing titers (VNT_50S_) were calculated as the half-maximal inhibitory dose (ID_50_) using 4-parameter nonlinear regression (GraphPad Prism). An overview of the VNT assay and examples of cells treated with both variants from one vaccinated and one infected individual’s serum can be found as (**Extended Data Fig. 7**)

### ACE2 competition assay

Biotinylated recombinant human ACE2 (#10108-H08H-B, Sino Biological) was diluted to a final concentration of 571.4 ng/mL in assay buffer (1:4 Low Cross Buffer (Candor Bioscience GmBH) in CBS (1X PBS + 1% BSA) + 0.05% Tween20) to create ACE2 buffer. Plasma samples were diluted 1:50 in assay buffer and then to a final concentration of 1:400 in ACE2 buffer under a sterile workbench, resulting in a final ACE2 concentration of 500 ng/mL in all samples. 25 μL of diluted plasma samples were then added to 25 μL of 1X BeadMix ^33^ per well of a 96-well plate (#3642, Corning). In addition to the standard bead mix used in MULTICOV-AB, all bead coupled RBD mutants were included. As a control, 500 ng/mL ACE2 was also used. Samples were incubated on at 21°C, 750 rpm for 2 hours on a Thermomixer. Samples were then washed using a microplate washer with Wash Buffer (1X PBS + 0.05% Tween20). 30 μL of 2 ug/mL Streptavidin-PE (#SAPE-001, Moss) was added to each well and incubated for 45 mins, 21°C, 750 rpm in darkness on a Thermomixer. Following incubation, samples were washed again and then resuspended in 100 μL of Wash Buffer, before being shaken again for 3 mins at 1,000 rpm. Samples were read individually on a FLEXMAP3D instrument under the following settings: 80 μL (no timeout), 50 events, Gate: 7,500-15,000, Reporter Gain: Standard PMT. MFI values were then normalized against the control wells. All samples were measured in duplicates and the mean is reported.

### Neutrobody Plex

The NeutrobodyPlex was performed as previously described ^35^ on all serum samples using a Nanobody concentration of 2 nM.

### Data Analysis

Data analysis and figure generation was performed in RStudio (Version 1.2.5001), running R (version 3.6.1) with the additional packages “beeswarm” and “RcolorBrewer” for data depiction purposes only. The type of statistical analysis performed (when appropriate) is listed in the figure legends. Figures were generated in Rstudio and then edited for clarity in Inkscape (Inkscape 0.92.4). Mann-Whitney U test was used to determine difference between signal distributions from different sample groups using the “wilcox.test” function from R’s “stats” library. Kendall’s τ coefficient was calculated in order to determine ordinal association between the observed antibody responses towards RBD mutant and wild-type proteins using the “cor” function from R’s “stats” library. Linear regression was performed to assess reduction in ACE2 neutralization observed for RBD mutants compared to wild-type proteins using the “lm” function from R’s “stats” library. Pre-processing of data such as matching sample metadata and collecting results from multiple assay runs was performed in Excel 2016. GraphPad Prism version 8.4.0 was used to process VNT data.

## Data Availability

The original data, figure files and data analysis scripts are available upon request.

## Data availability

Data relating to the findings of this study are available from the corresponding authors upon request.

## Code availability

Custom analysis code in R and required input files are available from the corresponding authors upon request.

## Author Contributions

UR, MK, MB, AD and NSM conceived the study. MB, AD, JaH, RF, AK, MiS, UR and NSM designed the experiments. KSL, MoS, GK, MiS, MK and NSM procured funding. MB, AD, DJ, NR, CH, JuH and ML performed experiments. TRP, MM, VM, TB, KA, RF, AK, MK, UR and NSM collected samples or organized their collection. PDK, BT and TW produced the RBD mutants. HP, MB and DJ produced and analyzed the QC samples in MULTICOV-AB. MB, AD, NR and MiS performed the data analysis. MB, AD, DJ and NSM generated the figures. AD wrote the first draft of the manuscript with input from MB, DJ, JaH, KSL, MoS, RF, AK, MiS, UR and NSM. All authors approved the final version of the manuscript.

## Acknowledgements

We thank Johanna Griesbaum for technical assistance, Florian Krammer for providing us with expression plasmids for the Spike Trimer and RBD, Tina Ganzenmüller and the diagnostic department of the Institute for Medical Virology, University Hospital Tübingen for providing patient samples and the NGS competence center of the University Hospital Tübingen for rapid sequencing. This work was financially supported by the State Ministry of Baden-Württemberg for Economic Affairs, Labour and Housing Construction (grant numbers FKZ 3-4332.62-NMI-67 and FKZ 3-4332.62-NMI-68), the Ministerium für Wissenschaft und Kunst Baden-Württemberg, the Initiative and Networking Fund of the Helmholtz Association of German Research Centres (grant number SO-96), the Deutsche Forschungsgemeinschaft (DFG-KO 3884/5-1) and the EU Horizon 2020 research and innovation programme (grant agreement number 101003480 – CORESMA). The funders had no role in study design, data collection, data analysis or the decision to publish.

---

**Extended Data Table 1.** Characteristics of Vaccinated and Infected sera.

| Characteristic | Vaccinated | Infected |
| --- | --- | --- |
| Number of donors | 23 | 35 |
| Median age (IQR) – years | 42 (16) | 59 (19) |
| Female sex (%) | 5 (21.7) | 10 (28.6) |
| 1<sup>st</sup> sample collection – median $\Delta T$<br>post first vaccination (range) | 21 days (-1 to 22 days) | Not applicable |
| 2<sup>nd</sup> sample collection – median $\Delta T$<br>post second vaccination (range) | 15 days (-7 to 16 days) | Not applicable |
| Median $\Delta T$ post positive PCR test<br>(range) | Not applicable | 14 days (2 – 85 days) |

For both vaccinated and infected groups, the number of donors, median age in years (including inter-quartile range) and number of females are provided. For vaccinated groups, the median time post-first vaccination is provided for the first sample, and the median time post-second vaccination is provided for the second sample. The full range is included for both samples. A total of 45 samples from 23 vaccinated donors were included in this study. For one individual a second sample was not taken. One individual had their first sample collected before they received a vaccination and so they have a negative $\Delta T$ for the first sample. Similarly, two individuals had their second sample collected before they received their second vaccination and so they have a negative $\Delta T$ for the second sample. For the infected group, the median $\Delta T$ for time between positive PCR test and sample collection is given including the full range.

**Extended Data Table 2.**
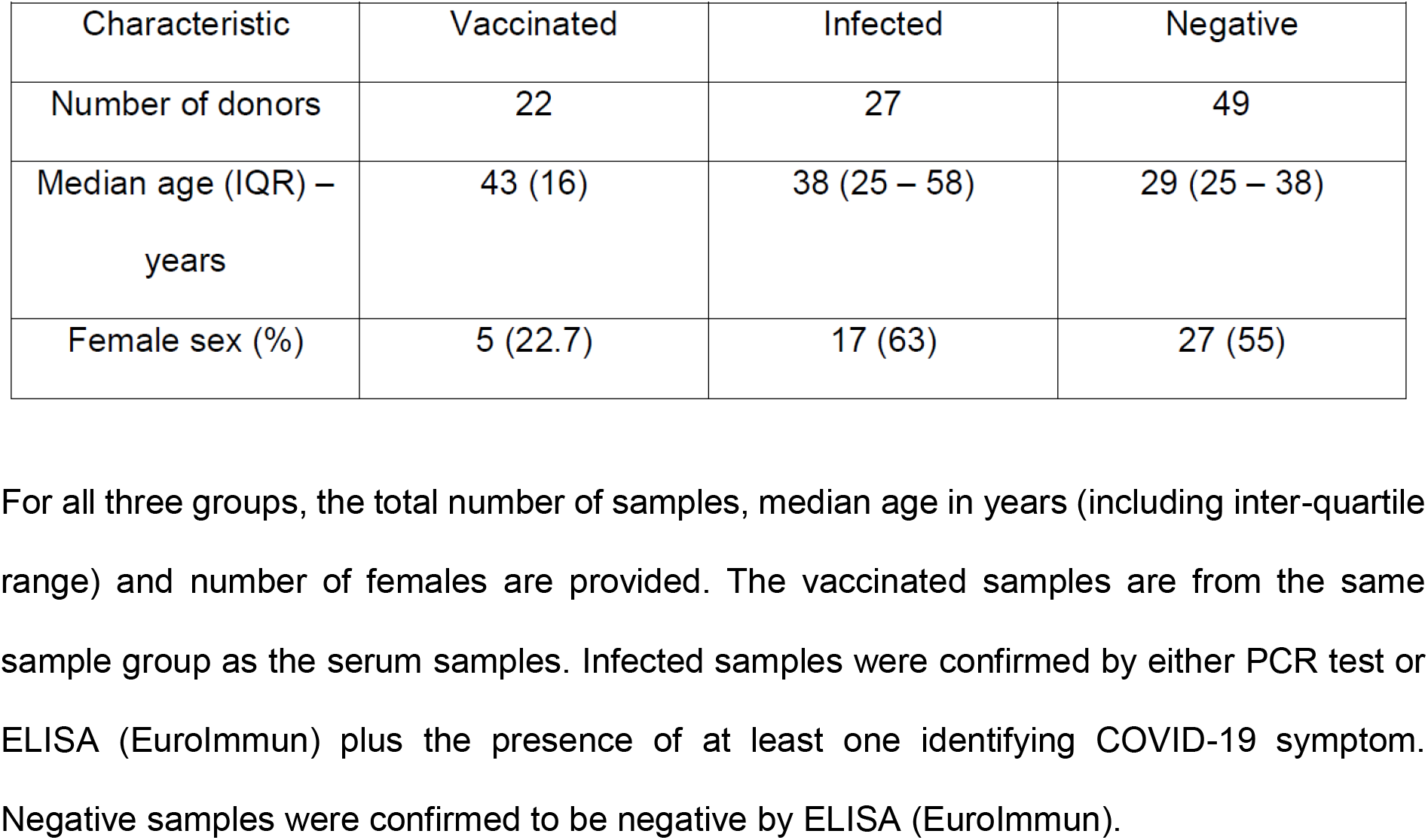
Characteristics of Vaccinated, Infected and Negative saliva samples.

**Extended Data Table 3.** Antigens included in MULTICOV-AB.

| Disease | Antigen | Manufacturer | Category number |
| --- | --- | --- | --- |
| SARS-CoV-2 | Spike Trimer | NMI | - |
| SARS-CoV-2 | RBD | NMI | - |
| SARS-CoV-2 | S1 domain | NMI | - |
| SARS-CoV-2 | S2 domain | Sino | 40590 |
| SARS-CoV-2 | Nucleocapsid | Aalto | 6404-b |
| SARS-CoV-2 | Nucleocapsid N-terminal domain | NMI | - |
| hCoV-OC43 | Spike | Sino | 40607-V08B |
| hCoV-OC43 | S1 domain | NMI | - |
| hCoV-OC43 | Nucleocapsid | NMI | - |
| hCoV-OC43 | Nucleocapsid N-terminal domain | NMI | - |
| hCoV-HKU1 | S1 domain | NMI | - |
| hCoV-HKU1 | Nucleocapsid | NMI | - |
| hCoV-HKU1 | Nucleocapsid N-terminal domain | NMI | - |
| hCoV-NL63 | Spike Trimer | NMI | - |
| hCoV-NL63 | S1 domain | NMI | - |
| hCoV-NL63 | Nucleocapsid | NMI | - |
| hCoV-NL63 | Nucleocapsid N-terminal domain | NMI | - |
| hCoV-229E | S1 domain | NMI | - |
| hCoV-229E | Nucleocapsid | NMI | - |
| hCoV-229E | Nucleocapsid N-terminal domain | NMI | - |

Full list of antigens included as standard in MULTICOV-AB (minus controls), their manufacturer, and if available, their category number. Full details on all NMI produced antigens can be found in <sup>33</sup>.

**Extended Data Figure 1.**
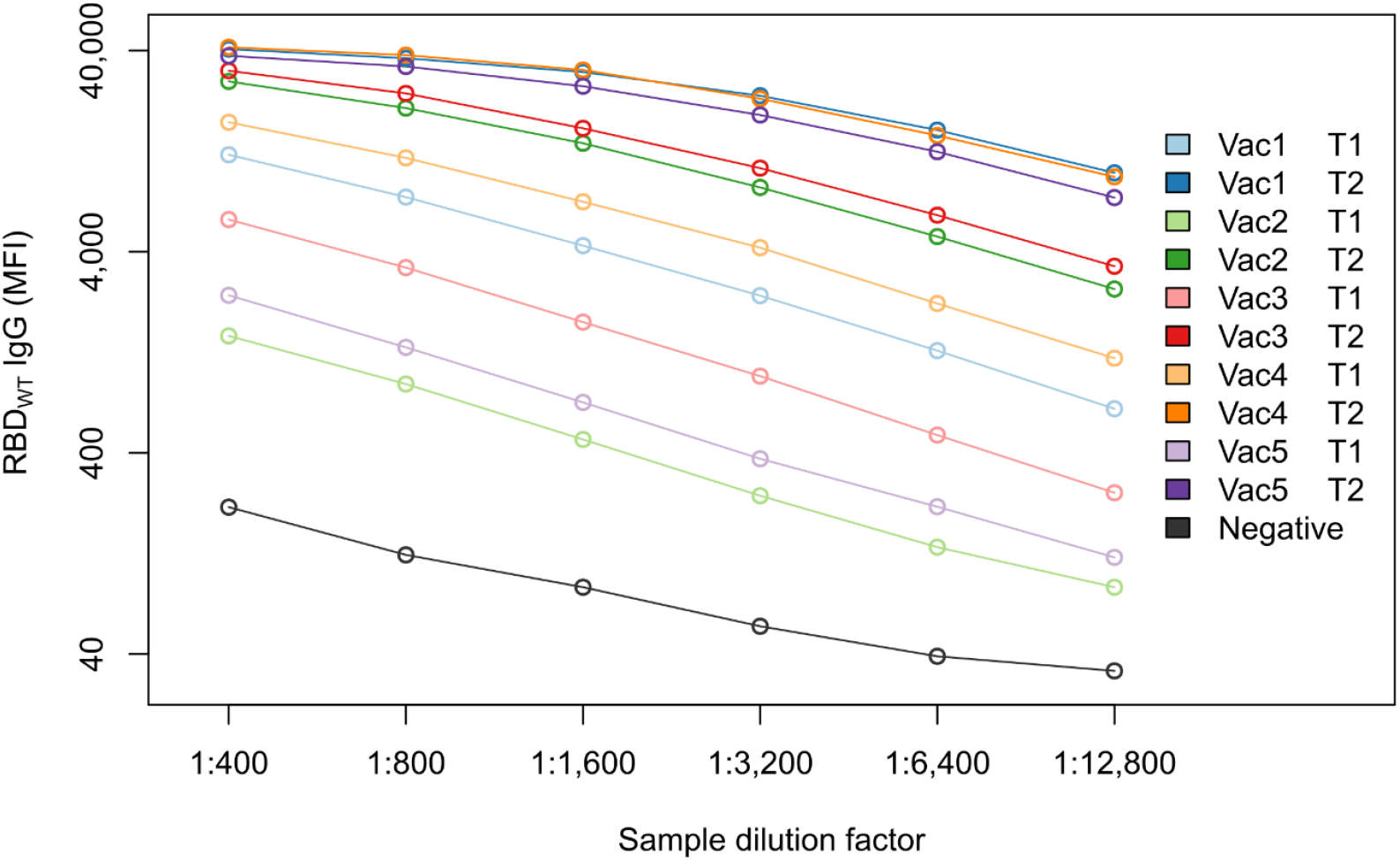
High serum antibody titers after the second vaccination dose. The second vaccination appears to plateau the serum antibody response at the Upper Limit of Detection for MULTICOV-AB. To confirm this, 11 samples consisting of 10 paired samples from five vaccinated individuals and one negative individual were examined in a dilution series. The three sera with the highest response maintained a similarly high response for the initial dilution, indicating a plateau in the range of >40,000 MFI, therefore suggesting that even for donors with high responses in the first sample, the second vaccination strongly increased the antibody response. A uniform curve shape for all samples (including the negative control) confirmed reliability of the generated data.

**Extended Data Figure 2.**
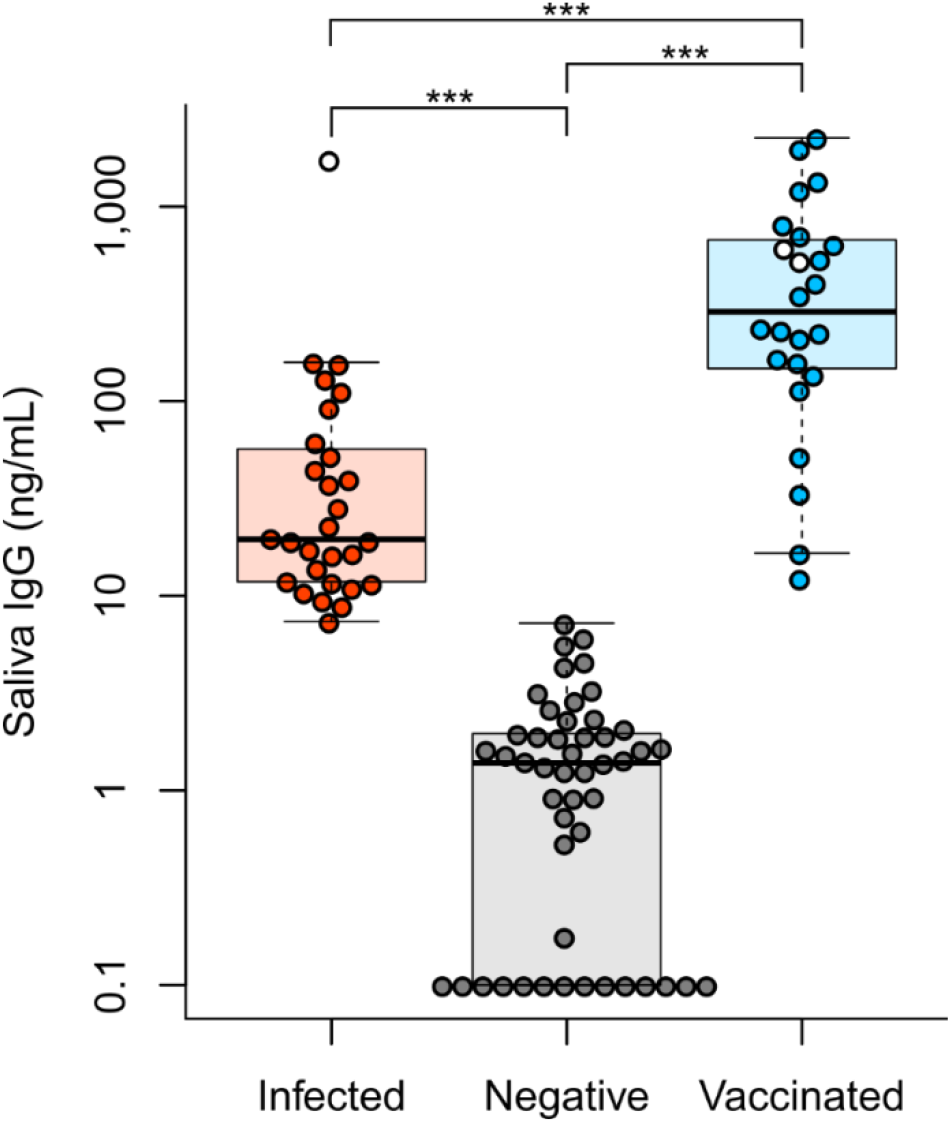
RBD reactive IgG detected by ELISA. To confirm results for MULTICOV-AB, anti-RBD IgG antibodies in saliva were measured by an in-house ELISA. As in Fig 2, saliva IgG response was highest in vaccinated individuals. Results are displayed as a Box and whisker plot (1.5 IQR). Two vaccinated sera samples from individuals not in contact with active SARS-CoV-2 infected individuals and one sera sample from an individual who was previously infected with SARS-CoV-2 and then later vaccinated are included as the blank circles. Negative samples that measured 0 were raised to 0.1 for display purposes only. For statistical analysis, their true value was used. Mann-Whitney U was used to determine statistical significance between the groups. *** indicates p-values lower than 0.0001.

**Extended Data Figure 3.**
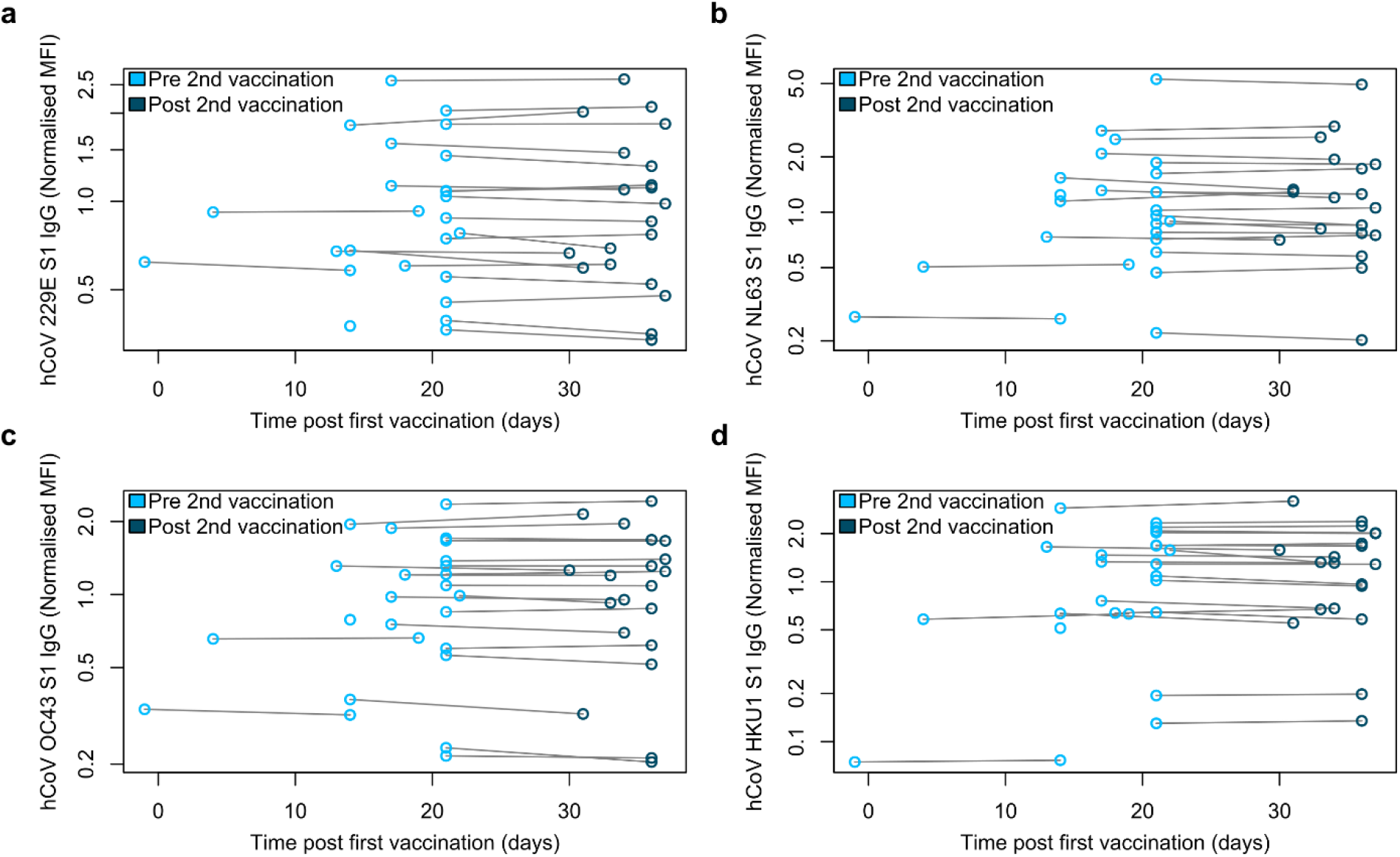
Cross-reactivity of antibodies to endemic coronaviruses in vaccinated individuals. Vaccinated individuals did not have an increased antibody response towards S1 proteins of the endemic coronaviruses 229E (**a**), NL63 (**b**), OC43 (**c**) and HKU1 (**d**). All samples were measured using MULTICOV-AB. Light blue (N=25) indicates samples are pre second vaccination, while dark blue (N=20) indicates samples are post second vaccination. Lines indicated paired samples from the same donor.

**Extended Data Figure 4.**
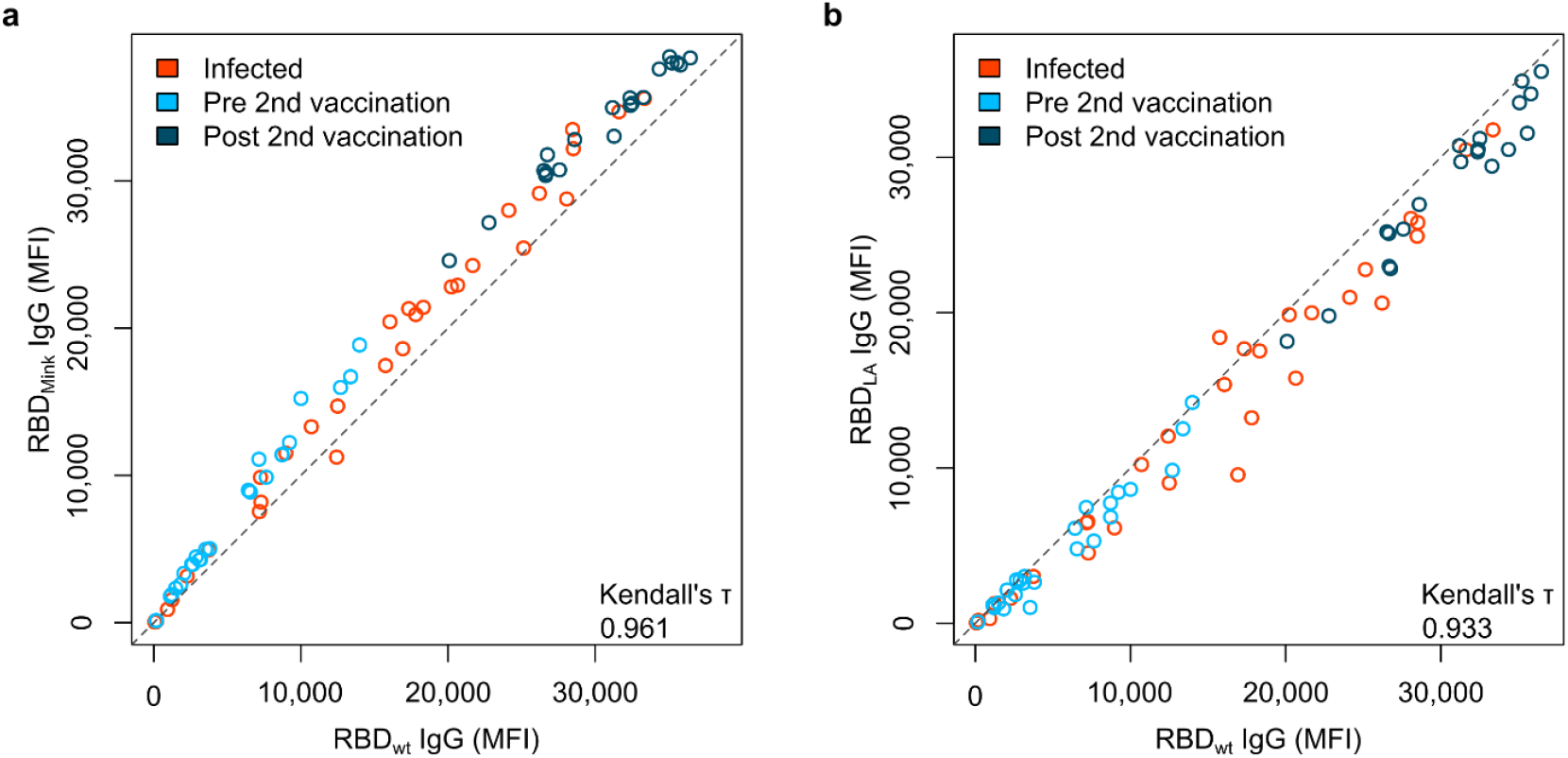
RBD mutants for the LA and Mink variants have similar antibody binding compared to the wild-type variant. When compared to wild-type (wt), RBD mutants for both the Mink (**a**) and LA (**b**) variants of concern resulted in similar response. RBD mutant antigens were generated (LA) or purchased (Mink) and added to MULTICOV-AB to measure the immune response towards them from vaccinated (N=45) and infected (N=35) sera, compared to the wild-type RBD. A linear curve (y=x) is shown as a dashed grey-line to indicate identical response between wild-type and mutant. Kendall’s tau was calculated to measure ordinal association between the mutant and wild-type.

**Extended Data Figure 5.**
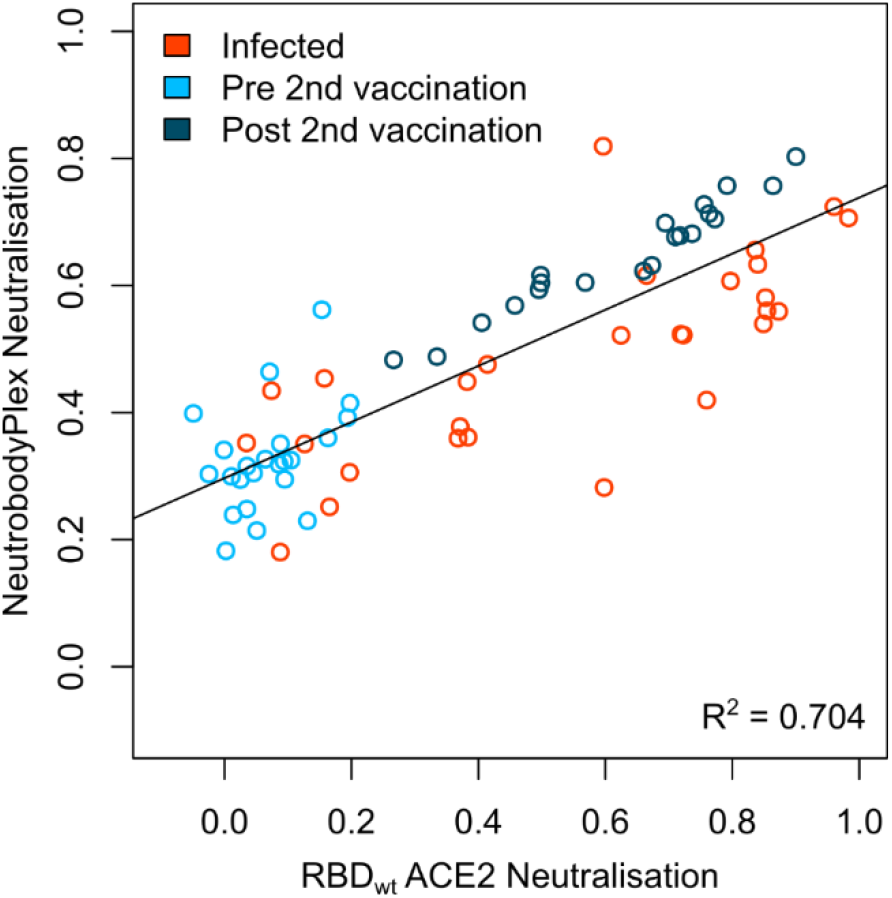
NeutrobodyPlex confirms reduced neutralization potential of vaccinated and infected sera for the wild-type variant. To further validate the results of the VNT and ACE2 competition assay, NeutrobodyPlex was used to examine neutralization potential of sera from vaccinated (pre-second dose (light blue, N=23), post-second dose (dark blue, N=20)) and infected individuals (red, N=28) for wild-type (wt) RBD. The correlation between NeutrobodyPlex and ACE2 competition assay results is shown. A linear regression (y=x) was calculated with the R^2^ value shown.

**Extended Data Figure 6.**
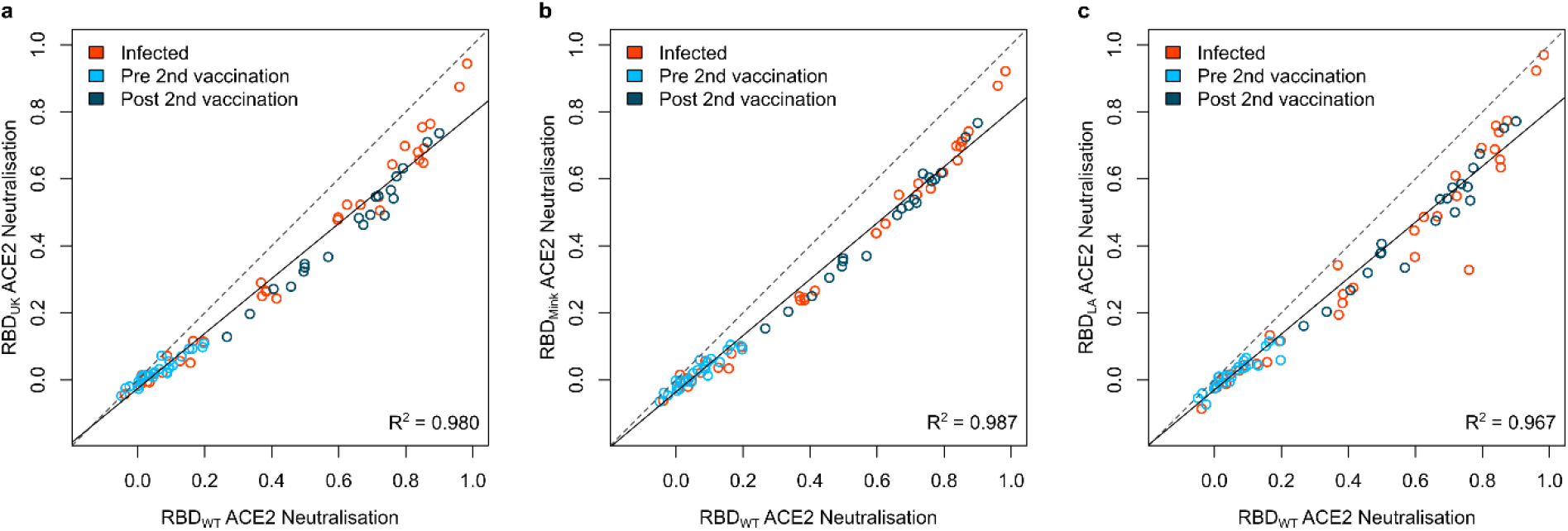
Neutralization as measured by ACE2 inhibition assay for the UK, Mink and LA RBD mutants compared to wild-type RBD. To determine the effect variants of concern had upon neutralization potential, an ACE2 competition assay was developed. RBD mutants for all variants of concern included within this manuscript (UK (**a**), Mink (**b**), LA (**c**)) were examined as well as the wild-type (wt) variant on sera from infected (red, N=35) and vaccinated (pre second vaccination (light blue, N=25), post second vaccination (dark blue, N=20)) individuals. Linear regression (y=x) was calculated for each panel, with the R^2^ value shown. Linear regressions had the following equations for the different figure panels: (**a**) y = −0.026 + 0.820x (**b**) y = −0.036 + 0.840x (**c**) y = −0.03 + 0.835x

**Extended Data Figure 7.**
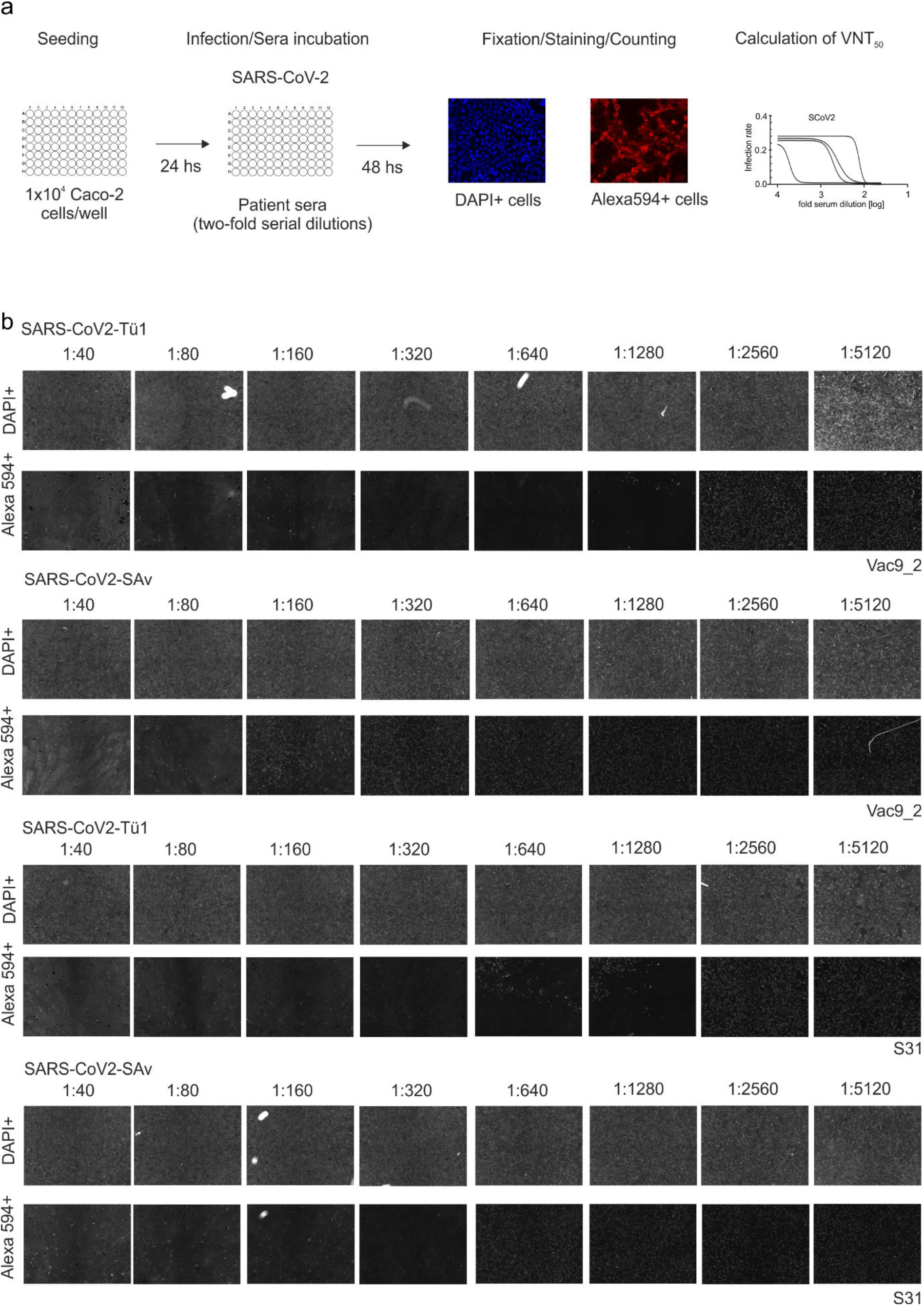
Overview of VNT. Simple overview of the VNT protocol (**a**) with examples (**b**) for cells treated with the wild-type (SARS-CoV-2-Tü1) and South African (SARS-CoV-2-SAv) for sera from one vaccinated (Vac9_2) and one infected (S31) individual. A dilution series (1:40 – 1:5120) is shown with the corresponding images for DAPI-positive and Alexa 594-positive cells.

## Notes

### Funding Statement

This work was financially supported by the State Ministry of Baden-Wuerttemberg for Economic Affairs, Labour and Housing Construction (grant numbers FKZ 3-4332.62-NMI-67 and FKZ 3-4332.62-NMI-68), the Ministerium for Wissenschaft und Kunst Baden-Wuerttemberg, the Initiative and Networking Fund of the Helmholtz Association of German Research Centres (grant number SO-96), the Deutsche Forschungsgemeinschaft (DFG-KO 3884/5-1) and the EU Horizon 2020 research and innovation programme (grant agreement number 101003480-CORESMA). The funders had no role in study design, data collection, data analysis or the decision to publish.

### Author Declarations

This study was approved the Ethics Committee of Eberhard Karls University and the University Hospital Tuebingen under the approval number 312/2020BO1 (Coro-Buddy) to Dr. Andrea Kreidenweiss, Institute for Tropical Medicine, University Hospital Tuebingen and Eberhard Karls University Tuebingen, and 312/2020BO2 to Dr. Karina Althaus, Institute for Clinical and Experimental Transfusion Medicine, University Hospital Tuebingen . All participants gave written informed consent.

